# Potentially modifiable mediators of the association between child abuse and dementia

**DOI:** 10.64898/2026.07.07.26357433

**Authors:** Katherine Taylor, Laura D Howe, Rebecca E Lacey, Emma Anderson, Naaheed Mukadam

## Abstract

**Background:** Literature investigating mediation of the association between child abuse and dementia has largely considered composite adverse childhood experience scores rather than individual adverse experiences, despite evidence that different experiences have different impacts on dementia risk. Additionally, prior studies consider mediators in isolation, despite known associations between mediators which may impact indirect pathways from child abuse to dementia.

**Objectives:** To investigate whether potentially modifiable health and lifestyle factors mediate the association between child abuse and dementia.

**Methods:** We used data from the English Longitudinal Study of Ageing to investigate associations between child abuse and dementia (N:5,448). Indirect pathways through four mediator categories (education, health behaviours, mental health and cardiovascular health) were examined. We used regression modelling to estimate associations between child abuse, mediators and dementia, and causal mediation analysis using the g-formula to estimate the joint indirect effect through the mediators.

**Results:** Individuals who experienced child abuse had, on average, an 80% higher hazard of dementia, compared to those who did not (RTE HR:1.80, 95% CI:1.21-2.39). Mental health mediators showed strong associations with both child abuse and dementia. Evidence for other mediators was weaker. Education, health behaviours, mental health and cardiovascular health mediated approximately 18% of the association. Sensitivity analysis revealed that almost all this mediation occurred through mental health.

**Conclusions:** Child abuse was associated with higher risk of dementia. Joint mediation analysis suggested that education, health behaviours, cardiovascular health, and mental health accounted for a relatively small proportion of the observed association, with most mediation occurring through mental health. Future research must focus on other potential pathways from child abuse to dementia, including biological and social mechanisms.

**Highlights:**

- On average, child abuse was associated with an 80% higher hazard of dementia.
- Modifiable health and lifestyle factors explained only 18% of the association.
- Mental health accounted for most of the observed mediation.

## Introduction

Child abuse has life-long health impacts and was previously observed to be associated with a 74% increased hazard of dementia (1,2). Unfortunately, child abuse cannot always be prevented. Although our focus should be primary prevention, understanding the causal pathways linking child abuse to dementia could inform the design of mitigation interventions for secondary prevention amongst individuals exposed to child abuse.

Most literature investigating mediation of the association between child abuse and dementia uses data from the UK Biobank (UKB), a large cohort study. A subset of participants completed an online mental health questionnaire where Adverse Childhood Experiences (ACE) were self-reported (N∼157,000). Associations between dementia and abuse were found to be partially explained by socioeconomic, behavioural, and medical factors. These factors mediated 26%, 24% and 16% of the association between dementia and emotional abuse, physical abuse and sexual abuse respectively (3). Most existing studies investigate each mediator in isolation, not considering the potential for pathways through multiple mediators. Multiple factors including depression, smoking, hypertension and diabetes were observed to mediate a small proportion of the association (4–6). One study found that post-traumatic stress disorder (PTSD) symptoms, but not diagnosis, mediated 75.3% of the association. Sub-diagnostic symptoms may influence the association, or the measure used may have lacked the discriminant validity to distinguish between PTSD and generalised distress (6).

Literature using other data sources is more limited. A study (N=11,601) in the China Health and Retirement Longitudinal Study (CHARLS), a cohort study of Chinese residents aged over 45, found depression to mediate 34.3% of the association between ACE and dementia. They found no evidence of mediation by smoking, sleep, alcohol, adult adverse experiences or adult socioeconomic status (SES)(7). Another study (N=8,953) using data from the Health and Retirement Study, a longitudinal panel study in the USA, found that years of education fully mediated the association. After considering education, there was limited evidence of a direct association between ACE and dementia. They also observed that the association was partially mediated by household income in midlife, but not physical activity in later-life (8).

Most studies focus on the association between dementia and ACEs, which consider abuse as one of many factors in an ACE sum score. ACE scores assume that all ACEs have the same impact on dementia risk. In prior work we observed greater variability in the impact of different ACEs on dementia risk than would be expected about a common hazard ratio. Only child abuse was strongly associated with increased dementia risk (2). Therefore, use of ACE scores as an exposure may dilute associations between dementia and child abuse, thereby diluting indirect associations through potential mediators.

ACEs, including child abuse, disproportionately affect those with low incomes. Therefore, it is important that studies investigating this association represent this population (9). However, UKB participants are healthier, more highly educated and of higher SES compared to the general UK population. Therefore, findings from the UKB may lack generalisability to the most vulnerable populations (10).

Existing studies often consider each mediator in isolation. Mediators do not occur independently of each other and the temporal sequence of events along pathways including multiple mediators is unclear. Most studies apply the counterfactual framework to decompose the total effect of ACEs on dementia into indirect and direct effects. However, this approach relies on strong assumptions whereby adjusting for intermediate confounders would introduce collider bias, but not adjusting for them violates the assumption of unmeasured confounding (11,12). Therefore, many of these studies may not satisfy the assumptions required by their methodologies.

We used data from the English Longitudinal Study of Ageing (ELSA), which is more representative of the UK population compared to UKB, to investigate mediation of the association between child abuse and dementia. By focussing on child abuse, we acknowledge that the pathways from different ACEs to dementia may differ and explore the mediators that may be specific to the association between child abuse and dementia. We estimate interventional effects for mediation analysis to consider the joint mediatory effect of four categories of potentially modifiable mediators, allowing pathways through multiple mediators. By identifying modifiable risk factors on the causal pathway from child abuse to dementia, and modelling the impact of intervening on them, we can evaluate whether joint interventions on health and lifestyle factors could potentially attenuate dementia risk amongst individuals who have experienced child abuse.

## Methods

### Study population

We obtained data from ELSA (www.elsa-project.ac.uk), a longitudinal panel study of English adults aged ≥50. The baseline interview took place in 2002-2003 and comprised a nationally representative sample of 11,391 individuals (mean age: 65.3, SD:10.4). The sample was refreshed 4 years later with new participants entering their 50s, to retain a structure representative of English adults aged over 50. Participants were interviewed every two years and had a health examination every four years.

### Sample selection

Participants within this study were restricted to those who have completed a retrospective life history interview (2007) and online mental health questionnaire containing information on child abuse. We restricted analyses to core study members, defined as age-eligible sample members interviewed at wave 1 or 3 of data collection. Participants with dementia at wave 3, or who retrospectively reported dementia diagnosis prior to their life history interview, were excluded as were those with no dementia data post wave 3 (Figure 1).

**Figure 1.**
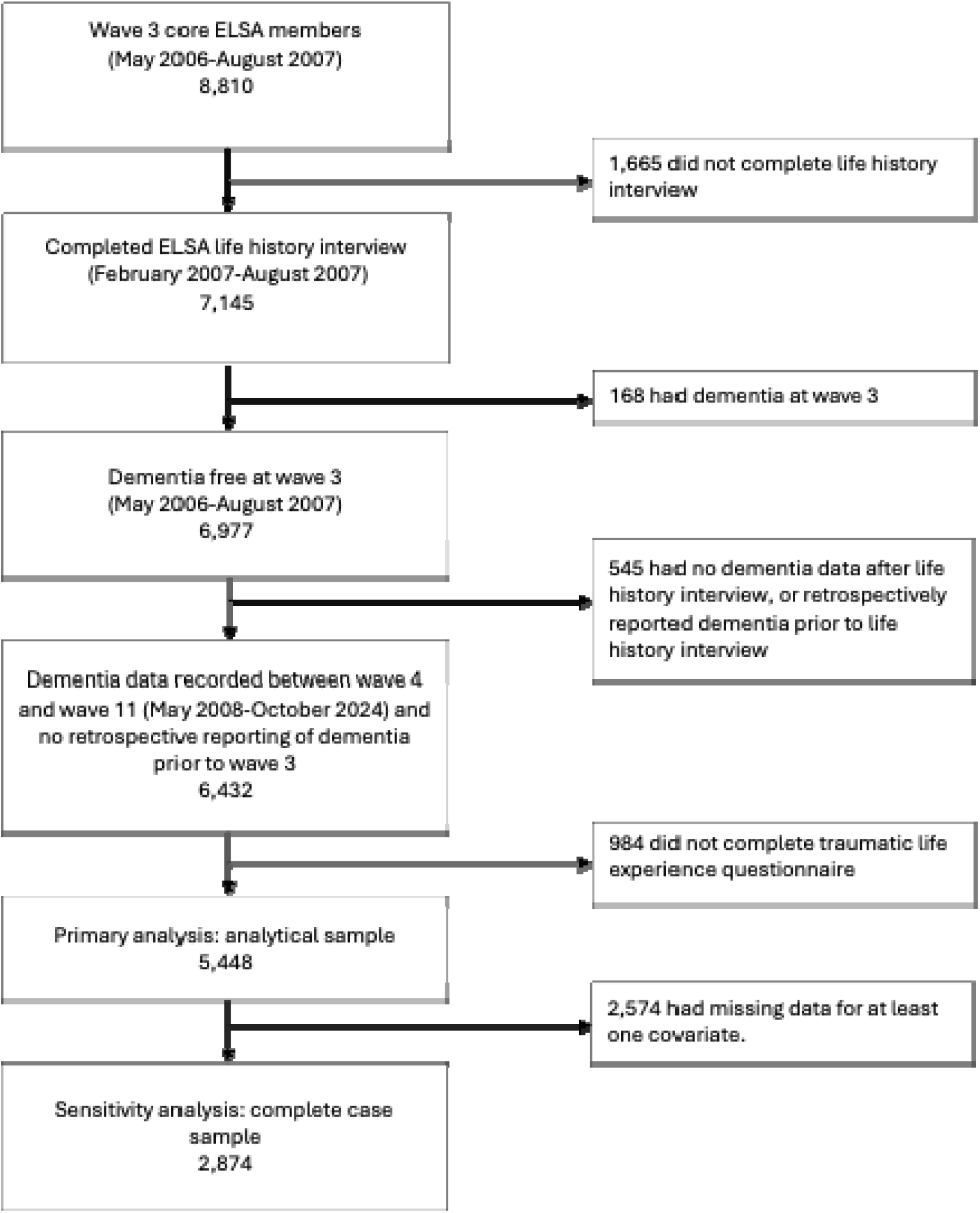
STROBE diagram of sample selection from core wave 3 ELSA members.

### Exposure

Child abuse was reported in the life history interview (average age 66.1 years, range 51-90+) and defined as having experienced at least one of the following whilst under the age of 16: sexual assault; serious physical attack or assault or having physically abusive parents.

### Outcome

We defined all-cause dementia algorithmically, in line with DSM-IV criteria as previously described (13,14). Briefly, dementia was defined as having self-reported doctor diagnosed dementia (N=284), scoring 3.6 or higher on the Informant Questionnaire on Cognitive Decline (N=32)(15) or having decline in two or more cognitive domains and a non-transient impairment in one or more activities of daily living (N=241). Time to dementia (months) was based on reported diagnosis date where available; otherwise derived from age at diagnosis or the midpoint between the first dementia report and last dementia-free interview. Follow-up began at the life history interview, with the earliest diagnosis date used for each participant.

### Mediators

We defined mediators as potentially modifiable risk factors which may lie on the causal pathway from child abuse to dementia (1,16,17). Mediators were selected based on the 2024 Lancet Commission on Dementia prevention, intervention, and care. Dementia risk factors were considered mediators if there was evidence linking them to child abuse. Anxiety was additionally included due to its established association with child abuse and mixed evidence as a dementia risk factor (18). We grouped mediators into four categories: cardiovascular risk factors (hypertension, obesity, diabetes, cholesterol); unfavourable health behaviours (smoking, alcohol consumption, physical inactivity); education (age leaving education) and mental health conditions (depression, anxiety).

Data on mediators was measured in ELSA wave 1 (2002-2003), wave 2 (2004-2005) or wave 3 (2006-2007). Physical measurements were taken by a nurse at wave 2. Self-reported health conditions, medication use and smoking status were obtained from harmonised variables which compiled data from all three waves. Self-reported health behaviour variables were measured at wave 3. If data on alcohol use was not available at wave 3, we used wave 2 data. If data were not available at wave 2 data from wave 1 was used.

We defined binary categories of mediators based on existing literature where possible (19,20). Diabetes was defined as self-reported doctor diagnosis, self-reported insulin injecting or having a HbA1c above 6.5%. Hypertension was defined as self-reported doctor diagnosis, reporting taking medication to lower blood pressure or a measured blood pressure of 140/90 mmHg. High cholesterol was defined as self-reported doctor diagnosis, reporting taking medication to lower cholesterol or a measured LDL of 3.36 mmol/l or more (19). Obesity was measured at the wave 2 nurse visit and defined according to World Health Organisation definitions (21). Health behaviours were self-reported. Smoking was defined as being a current smoker. Frequent alcohol consumption was defined as drinking 5 or more days a week. Physical inactivity was defined as never or rarely doing vigorous or moderate exercise. Mental health conditions were defined according to self-reported doctor diagnosis of anxiety or depression. Depression was also defined as scoring three or more on the Center for Epidemiologic Studies Depression Scale (CESD-8). Age finishing education was measured in each wave of ELSA and harmonised. The variable was treated as continuous and reverse coded so that lower levels of education were denoted by a higher number. Composite measures were developed for cardiovascular risk factors and unfavourable health behaviours by creating a sum score based on the binarized individual mediators. A binary variable for any experience of anxiety or depression was generated.

### Confounders

Age and sex were self-reported at the life history interview (2007). Ethnicity was reported at wave 3 (2006-2007) and was binarized into White or Other ethnicities due to low sample numbers for the latter. Parental occupation at age 14 was measured in waves 1-3 and harmonised. We coded managers, senior officials, business owners, professionals, and technicians as highly skilled. Participants employed in the armed forces were coded as missing (N=142) due to lack of clarity about the nature of the work (22). All other categories were coded as lower skilled (9).Three additional proxy measures for childhood SES at age 10 were measured at the life history interview (23). Number of books was binarized into ten books or fewer, and more than ten books. Number of bedrooms per capita and number of facilities in accommodation were used to create two continuous variables. In all model’s childhood SES proxies were added individually.

### Statistical analysis

We conducted analyses in RStudio (2025.09.0+387) and examined missing data patterns using regression models. Multiple imputation by chained equations was applied to 2,574 participants, and imputed data used for our primary analysis (Figure 1). The imputation model included child abuse measures, mediators, age, sex, childhood SES, ethnicity, dementia incidence, time to dementia, and censoring by wave 10, with income, education, occupation, and broader adverse experiences as auxiliary variables. Number of imputations was determined using a two-stage approach. Our initial pilot generated 20 datasets (10 iterations), and Monte Carlo error was assessed for key regression coefficients, with <10% of the standard error deemed acceptable. Based on this, we generated 40 datasets (10 iterations) for the final analysis (24,25). Composite mediators were derived post-imputation. Sensitivity analyses were conducted using complete case data. Sample characteristics were described and variables cross-tabulated by experience of child abuse and dementia incidence.

Theoretically, a mediator on the causal pathway from child abuse to dementia should be associated with both child abuse and dementia. We used logistic regression models to estimate associations between child abuse and binary mediators. Linear regression models estimated associations between child abuse and continuous mediators. Cox proportional hazards models estimated associations between all mediators and dementia free survival time. We evaluated the proportional hazards assumption by testing for time-dependent effects of covariates via scaled Schoenfeld residuals (26). All models were adjusted for sex, age, age squared, and childhood socioeconomic status.

Figure 2 shows the proposed causal pathway from child abuse to dementia. We assessed mediation using interventional effects to examine pathways through mental health, cardiovascular health, education, and health behaviours, with composite scores representing each mediator domain. Cox proportional hazards models were used to estimate associations between child abuse and dementia-free survival, as well as between each mediator and dementia-free survival. Logistic regression was applied to binary mediators (anxiety and depression), and linear regression to continuous mediators (cardiovascular score, health behaviour score, and years of education). We adjusted for sex, age, age squared, and childhood socioeconomic status in all models.

**Figure 2.**
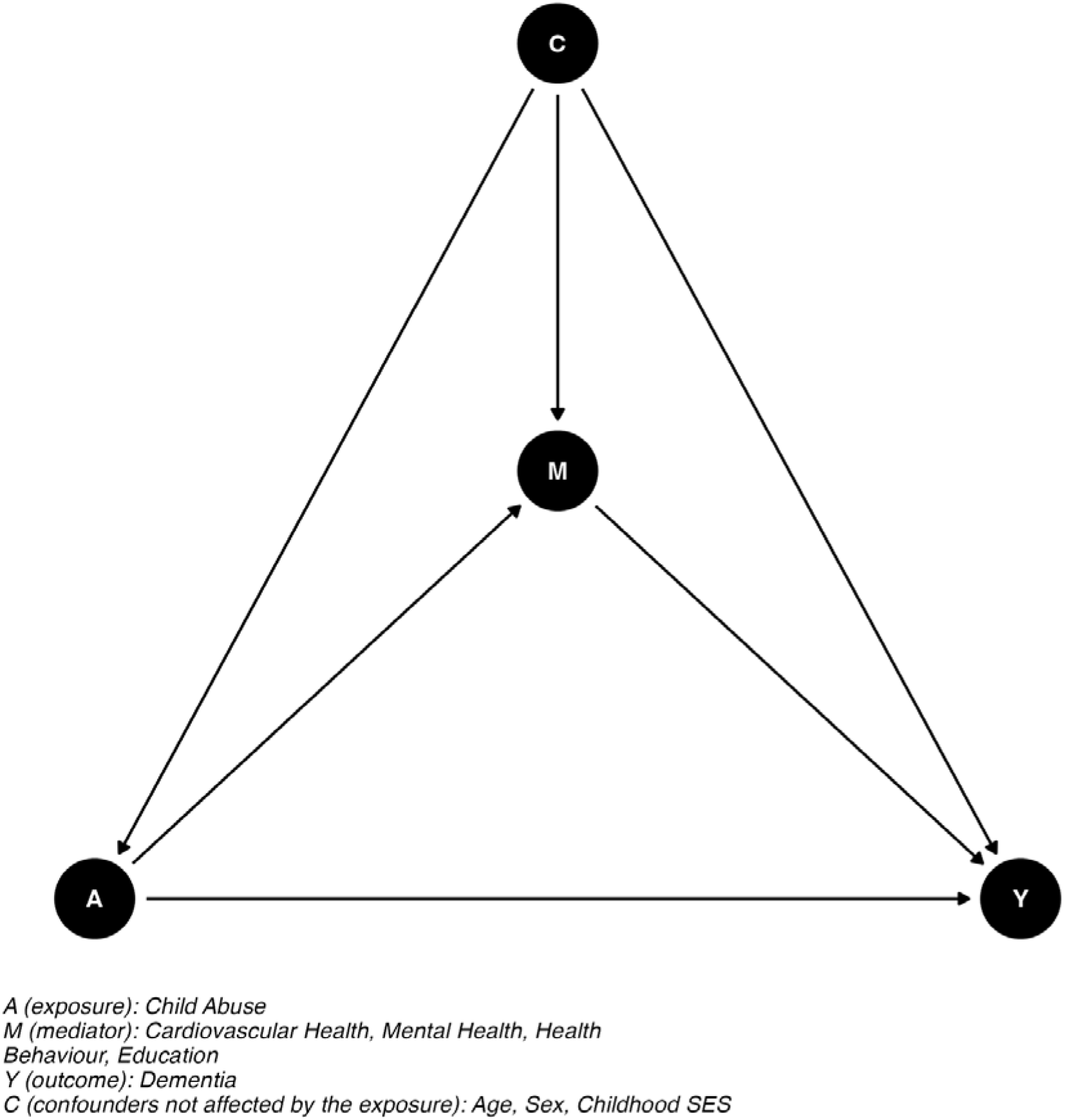
Directed Acyclic Graph of mediation analysis.

Using the g-formula, we modelled the impact of jointly intervening on the distribution of all four mediators (27). This approach was selected because our chosen mediators are not independent of each other and their temporal ordering could not be established, making single-mediator analyses likely to violate causal mediation assumptions (11,12). The estimation procedure has been described elsewhere (11,28,29). Briefly, this approach works by modelling the impact of jointly intervening on the distribution of mediators and then calculating the impact of this distribution change on dementia risk among those exposed to child abuse. 95% CI were generated using 1000 bootstrap samples. We decomposed the association between child abuse and dementia into effects including: The Randomised Total Effect (RTE), The Randomised Natural Direct Effect (RNDE), The Randomised Natural Indirect Effect (RNIE) and Proportion Mediated (PM). The RTE can be interpreted as the total effect of child abuse on dementia. The RNDE can be interpreted as the effect of child abuse on dementia that is not mediated by the considered mediators. The RNIE represents the effect of child abuse on dementia that is mediated by the considered mediators. The Proportion Mediated (PM) reflects the proportion of the total effect estimated to operate through the considered mediators. It was calculated using the formula 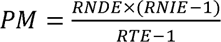.

### Sensitivity analysis

Childhood SES may have been measured after an individual’s first experience of abuse. Therefore, we ran all regression and mediation analysis models without adjustment for childhood SES. Given the results of regression models, we re-ran the causal mediation analysis including only mental health as a mediator. We ran the model once adjusting for age, sex and childhood SES measures and once also adjusting for other mediators as confounders. Primary analyses were repeated using complete case data. Regression models considering only LDL reported at wave 2 were conducted because other cholesterol measures captured a broad definition of high cholesterol. We used inverse probability weighting to investigate the potential for selection bias to influence the association between child abuse and education.

Additional analysis of the association between child abuse and education was conducted, first excluding those who reported no education due to a low number of participants in this category, then excluding those who reported their first experience of abuse occurring after they had finished education.

## Results

Our analytical sample consisted of 5,448 individuals (Figure 1). Included individuals had higher levels of income and education, were more likely to have a managerial or professional occupation and more likely to have parents in a highly skilled occupation at age 14 compared to those not included (Supplementary Table 1).

7.4% of the sample experienced child abuse and 10.2% of individuals developed dementia during follow-up. Participants were followed for an average of 10.9 years (Supplementary Table 2). Participants had a mean of 1.6/4 cardiovascular health conditions and a mean health behaviour score of 1/3. 27.9% had anxiety or depression and 48.5% of participants finished their education aged 15 or under. Dementia incidence was higher among those who experienced child abuse (10.9% compared to 10.2%) (Supplementary Table 3).

### Regression modelling

Child abuse was consistently associated with negative mental health outcomes. Child abuse was associated with more than twice the odds of developing depression (OR:2.24, 95% CI:1.81-2.77) and almost triple the odds of anxiety (OR:2.90, 95% CI:2.16-3.89). When a composite measure was considered, child abuse was associated with 120% higher odds of having depression or anxiety (OR:2.20, 95% CI:1.78-2.72) (Figure 3, Supplementary Table 4). Poor mental health was also consistently associated with higher hazard of dementia. Depression, anxiety and any experience of depression or anxiety was associated with 58% (HR:1.58, 95% CI:1.32-1.90), 58% (HR:1.58, 95% CI:1.15-2.19) and 56% (HR:1.56, 95% CI:1.30-1.87) higher hazard of developing dementia respectively (Figure 3, Supplementary Table 4).

**Figure 3.**
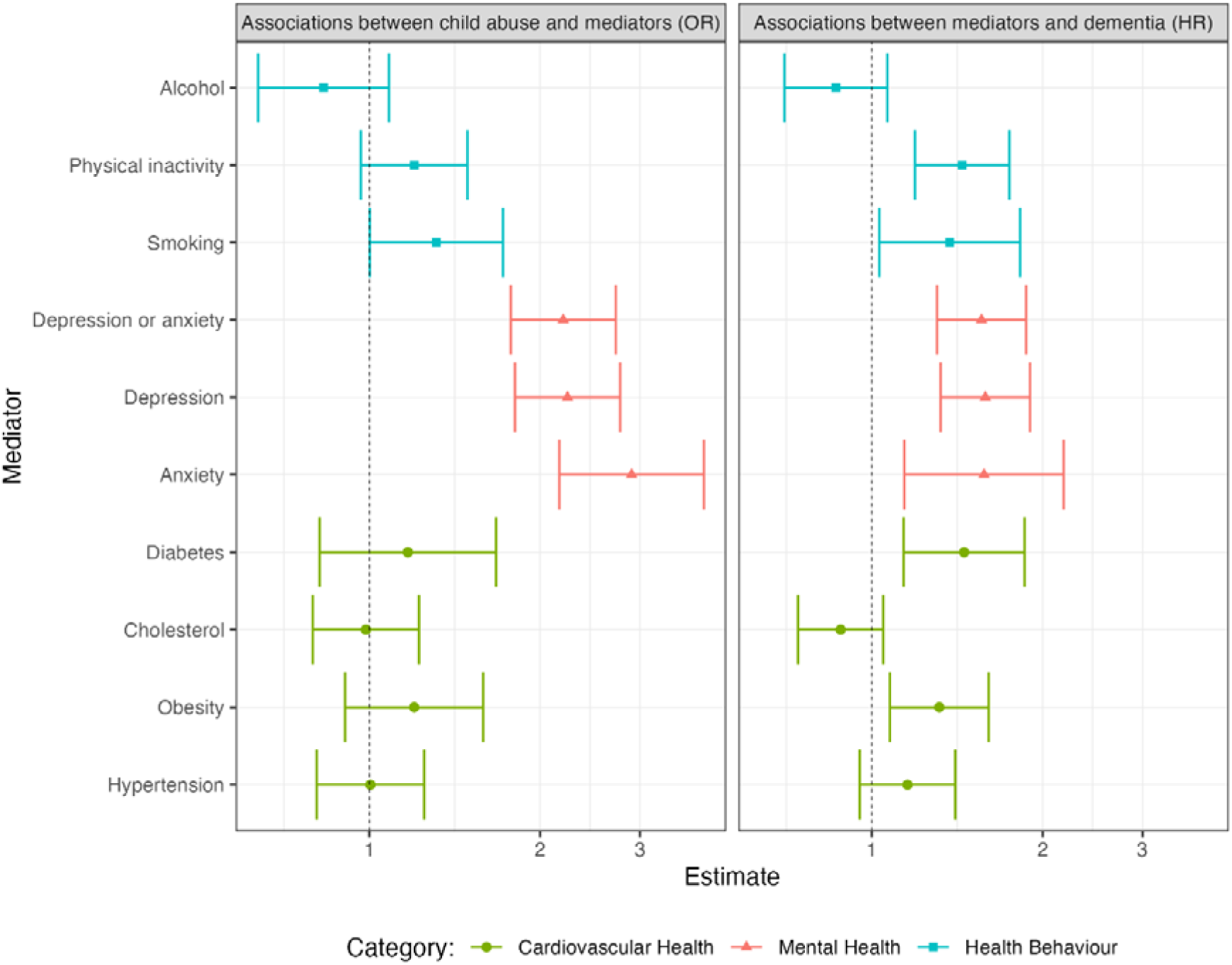
Regression models of the associations between exposure, binary mediators and outcomes. Logistic regression of mediators in association with child abuse. Cox proportional hazards modelling of dementia incidence in association with mediators.

We did not find strong evidence of associations between child abuse and health behaviour metrics. There was suggestive evidence of associations between child abuse and higher odds of smoking (OR:1.31, 95% CI:1.00-1.72), being physically inactive (OR:1.20, 95% CI:0.97-1.49) and lower odds of frequent alcohol consumption (OR:0.83, 95% CI:0.64-1.08). We found some evidence that child abuse was associated with a higher negative health behaviour score, but the estimate was imprecise (β:0.05, 95% CI: −0.02-0.12) (Supplementary Table 4). In primary analyses, all negative health behaviours aside from high alcohol consumption were associated with increased hazard of developing dementia (smoking HR:1.35, 95%.CI:1.02-1.80, physical inactivity HR:1.43, 95% CI:1.18-1.74, alcohol HR:0.88, 95% CI:0.71-1.08, health behaviour score HR:1.18, 95% CI:1.04-1.34).

We did not find strong evidence of associations between child abuse and cardiovascular health. Obesity (OR:1.20, 95% CI:0.91-1.59) and diabetes (OR:1.17, 95% CI:0.82-1.67) were positively associated with child abuse, but estimates were imprecise. We found little evidence of an association between child abuse and hypertension (OR:1.00, 95% CI:0.81-1.25) or cholesterol (OR:0.99, 95% CI:0.80-1.22) (Figure 3, Supplementary Table 4). There was weak evidence of an association between child abuse and a higher negative cardiovascular health score (β:0.05, 95% CI: −0.06-0.16). Hypertension (HR:1.15, 95% CI:0.95-1.40), obesity (HR:1.31, 95% CI:1.07-1.60) and diabetes (HR:1.45, 95% CI:1.13-1.85) were associated with a higher hazard of dementia (Figure 3, Supplementary Table 4) We found little evidence of an association between higher cholesterol and increased dementia risk (HR:0.88, 95% CI:0.74-1.05). Higher negative cardiovascular health score was associated with a higher hazard of developing dementia. Each additional cardiovascular health condition was associated with a 11% increase in dementia risk (HR:1.11, 95% CI:1.01-1.22).

We did not observe an association between child abuse and years of education (β: −0.01, 95% CI: −0.16-0.14) (Figure 4, Supplementary Table 4) nor between years of education and dementia incidence (HR:1.01, 95% CI:0.95-1.07) (Figure 4, Supplementary Figure 2, Supplementary Table 4).

**Figure 4.**
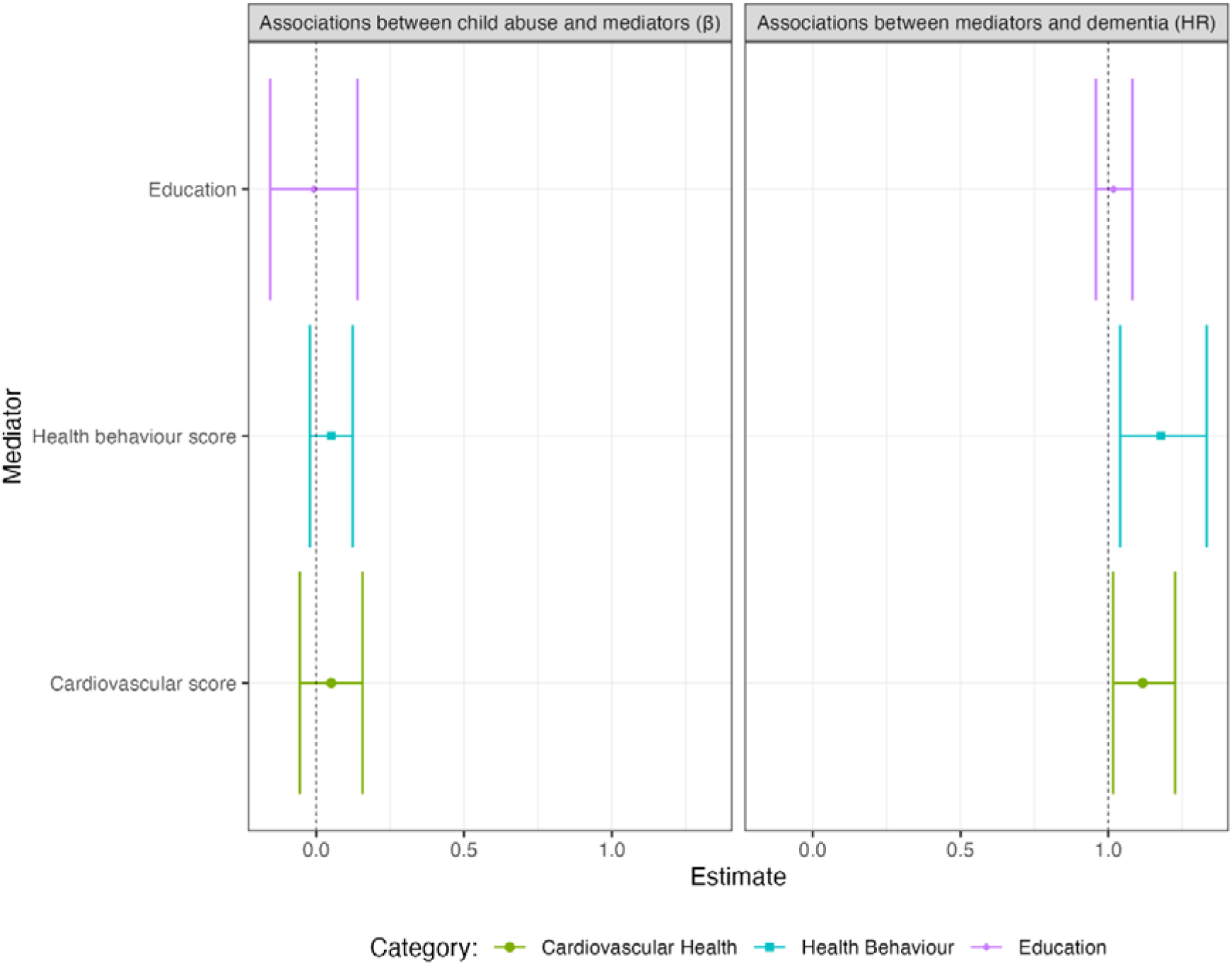
Regression models of the associations between exposure, continuous mediators and outcomes. Linear regression of mediators in association with child abuse. Cox proportional hazards modelling of dementia incidence in association with mediators.

All findings from the regression sensitivity analyses were consistent with the primary analyses (Supplementary Table 4-6, Supplementary Figure 1-2).

### Mediation analysis

In primary analysis, hazard of dementia was 80% higher amongst individuals who had experienced child abuse (RTE:1.80, 95% CI:1.21-2.39). Most of the association between child abuse and dementia was not mediated by years of education, health behaviours, mental health and cardiovascular health (RNDE:1.66, 95% CI:1.12-2.20). The RNDE was similar to the RTE indicating limited mediation of the association by the mediators considered.

Mediation by the considered factors was estimated to explain 18% of the association between child abuse and dementia. Jointly intervening on years of education, health behaviours, mental health and cardiovascular health amongst those who experienced child abuse was estimated to reduce their hazard of dementia by 8% (RNIE:1.08, 95 CI%:1.03-1.14). (Figure 6, Supplementary Table 7). Results from sensitivity analysis not adjusting for childhood SES were consistent with primary analysis. Estimates using complete case data were very imprecise and 95% CI for the total and direct effects of child abuse on dementia crossed the null (Supplementary Table 7, Supplementary Figure 3). In sensitivity analysis which only considered experience of depression or anxiety as a mediator, the RNIE was very similar to that for our primary analysis (RNIE:1.07, 95% CI:1.03-1.13). The proportion mediated by mental health was approximated as between 15 and 17% (Supplementary Table 7).

**Figure 6.**
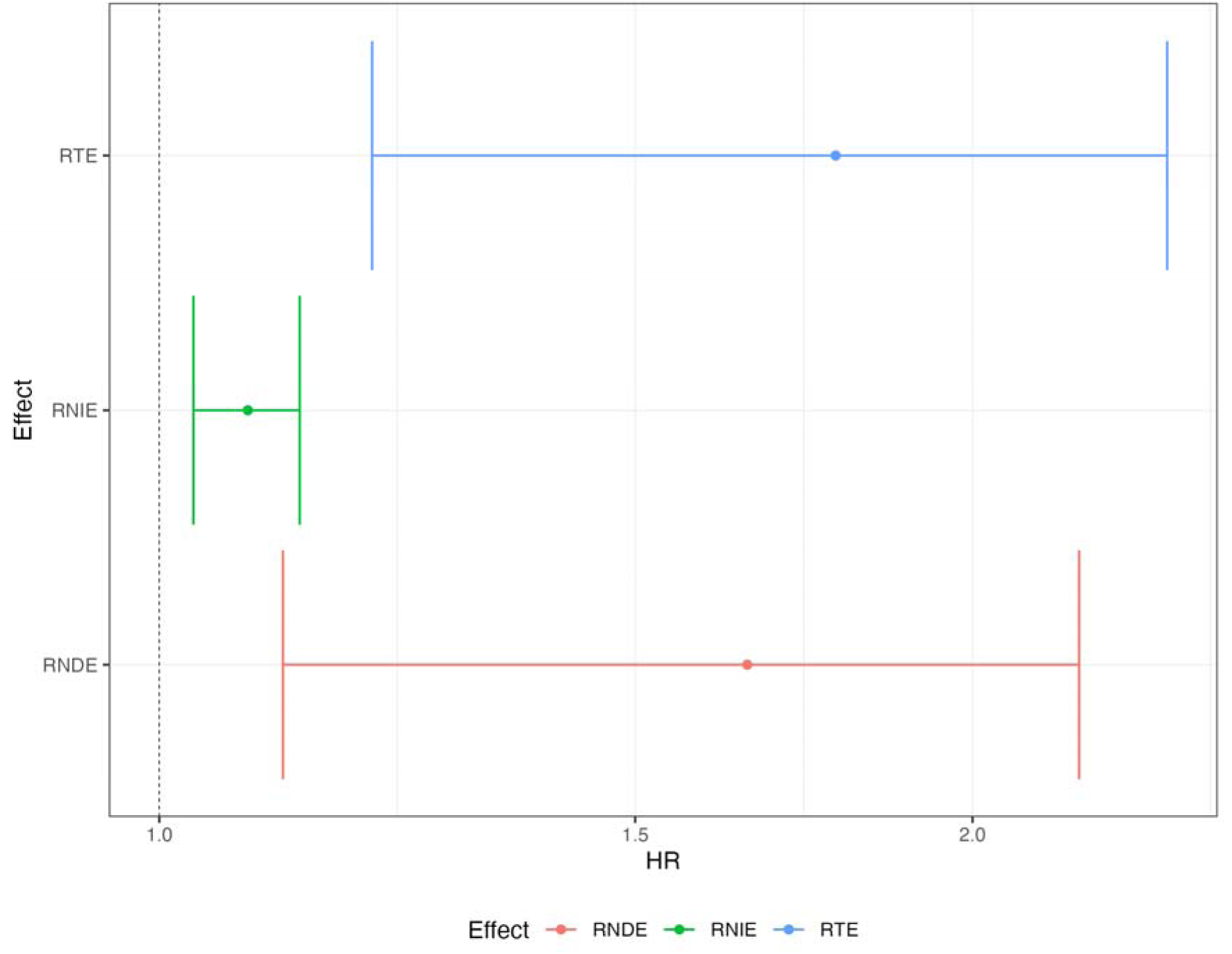
Randomised interventional effects for mediation analysis. Joint mediation effects of education, health behaviours, cardiovascular health and mental health on the association between child abuse and dementia incidence. RNDE: Randomised natural direct effect. RNIE: Randomised natural indirect effect. RTE: Randomised total effect.

## Discussion

### Key findings

Participants who experienced child abuse had, on average, an 80% increased hazard of developing dementia. Eighteen percent of this association was mediated by the potentially modifiable factors considered. Joint intervention on health behaviours, mental health, cardiovascular health and education amongst those who experienced child abuse was estimated to reduce their hazard of dementia by approximately 8%. Most of the observed mediation resulted from mental health and there was limited evidence of mediation by the other factors considered.

### Existing literature

Previous studies suggest that recognised modifiable dementia risk factors explain only a modest proportion of the association between ACEs and dementia, although estimates vary substantially depending on study population, analytical approach and adversity definition. We found that a modest proportion of the association between child abuse and dementia was mediated by health behaviours, mental health, cardiovascular health and education. A key contribution of our study is the estimation of interventional effects using the g-formula.

Unlike previous studies which examined mediators individually or used traditional mediation approaches, this novel framework estimates the joint contribution of multiple correlated mediators, without requiring as many strict assumptions which are unlikely to hold in complex causal systems (11,12).

One study in the UKB found that between 16% and 26% of excess risk of dementia associated with child abuse was mediated through sociodemographic factors, behavioural factors and medical history. This suggests that between 74% and 84% of the association occurs through alternative pathways, aligning with our findings (3). A broader study investigating the association between ACEs and dementia found 67% of the association was mediated through sociodemographic characteristics, social relationships, health behaviour and health status. The measure of ACE included a wider range of experiences which may influence dementia through more indirect pathways compared to child abuse (30).

Theoretically, to act as a mediator a variable should be associated with both the exposure and the outcome. In our analysis, only mental health was associated with both child abuse and dementia. Sensitivity analysis revealed that our mediation analysis estimates were predominantly driven by mental health (15-17% out of 18%). Within the UKB, depression is consistently found to mediate a small proportion, between 4.3% and 8.7%, of the association between ACEs and dementia (4–6). In CHARLS, the mediatory effect of depression was larger at 34.3% (7).

Existing literature has found limited evidence for mediation of the association between ACE and dementia through cardiovascular health. This aligns with our results although our observed associations between child abuse and cardiovascular health differ from some existing literature. One study observed no indirect effect from ACE to later-life cognition through systolic blood pressure (31). Another found no substantial mediation by chronic cardiovascular conditions (32). In the UKB only a very small proportion of the association between ACE and dementia was mediated by hypertension (1.8%), diabetes (2.6%), dyslipidaemia (1.3%) and stroke (1.5%). Additionally, there may be some overlap in the proportion mediated by each of these factors as each was considered in isolation (4).

Our findings provide limited support for health behaviours as mediators on the pathway from child abuse to dementia. We found suggestive evidence that child abuse was associated with a higher incidence of smoking, physical inactivity and a higher negative health behaviour score. Negative health behaviours, aside from alcohol consumption, were associated with increased incidence of dementia. Contrary to our expectations, child abuse associated with lower alcohol consumption. Within ELSA, only the frequency of alcohol consumption could be obtained which may contribute to our findings. Aligning with our results, a prospective cohort study from the United States also found that later-life physical activity did not mediate the association between ACE and dementia (8). Findings from studies in the UKB also suggest a limited mediatory role for smoking and alcohol consumption (4,5,7).

Unexpectedly, we did not observe an association between child abuse and education. This may reflect recall bias, those with higher levels of education may be less likely to report child abuse. Selection bias could have influenced our results, though inverse probability weighting did not change our findings. There is relatively little variation in age leaving education within our participants, alternatively our results may reflect cohort effects. Previous work in ELSA which conducted a path analysis from early-life adversity to later-life cognition also found no mediation through education whereas a similar analysis in UKB did (33). UKB participants had a lower mean age (58, SD:8.09) compared to ELSA participants (66, SD:9.58). The different sample structure may explain differences in the findings (33). Multiple studies find education to mediate the association between ACE and dementia or cognition. One study found education to mediate 23.2% of the association with intellectual function and 12.5% of the association with executive attention (34). A cohort study from the USA found education to fully mediate the association between ACE and dementia (8). However, both studies consider ACE and not abuse. Mediation through education may be more important for pathways from other adverse experiences to dementia.

### Potential mechanisms

Given we find modest evidence for mediation by our considered factors, other social or direct biological pathways may play an important role. Childhood adversity has wide ranging effects on neural, endocrine, immune and metabolic physiology (35). Several inflammation markers have been observed to mediate associations between ACE and dementia, though the proportion mediated was small (5). There are social mechanisms which we couldn’t capture using data from ELSA. ELSA does not measure PTSD symptoms, which in the UKB mediated a large proportion of the relationship (6). Child abuse is associated with a greater risk of later-life adversity, which is associated with heightened dementia risk (2,7). The structure of ELSA’s adverse experience questionnaire meant we could not consider adult adverse experiences as a mediator. Child abuse is associated with long-term poverty which is also linked to increased dementia risk via more complex pathways (19,36).

Our findings have important implications for dementia prevention. Current strategies focus on modifying cardiovascular risk factors and lifestyle behaviours in later-life. Whilst these remain important for reducing dementia incidence overall, our results suggest they are unlikely to substantially reduce the excess dementia risk experienced by individuals exposed to child abuse. Preventive strategies may need to incorporate earlier intervention, including trauma-informed mental healthcare and approaches targeting the long-term biological consequences of child abuse.

### Strengths and Limitations

ELSA is a well-characterised, nationally representative study, making our findings more generalisable to the older English population than some previous work. By focussing on the child abuse, rather than ACE, we consider that ACE have heterogenous impacts on dementia risk and may act through different indirect pathways. Use of interventional effects for mediation analysis, implemented using g-formula, enabled us to consider the joint effect of multiple mediators without violating the assumptions of causal mediation analysis. It also provides more policy specific interpretations compared to other methods. Additionally, we use multiple imputation and inverse probability weighting to account for missing data, attrition and selection bias.

Child abuse was measured retrospectively, and individuals may be reluctant to share details of their adverse experiences, resulting in underreporting. Many mediators were measured through self-report and so are subject to recall bias. Underreporting of both exposure and mediators may bias results toward the null. We could not measure the severity or timing of abuse which may result in us underestimating all examined associations. Causal mediation analyses rely on strong and untestable assumptions, including the absence of unmeasured confounding and a clear temporal sequence of events between exposure, mediators, and outcome. Although use of interventional effects implemented via the g-formula reduces methodological limitations, our results remain vulnerable to residual confounding, measurement error, and reverse causation. Some mediators, particularly depression and anxiety, may partially reflect prodromal dementia rather than causal intermediates and so mediation estimates should be interpreted cautiously.

### Conclusions

Individuals who experience childhood abuse are at increased risk of developing dementia, and a modest proportion of this association is likely to be mediated by cardiovascular health, mental health, health behaviours, and education, with most mediation being attributed to mental health. Identifying the biological and psychosocial mechanisms responsible for the remaining unexplained risk should be a priority for future research and may reveal novel opportunities for dementia prevention.

## Data availability statement

The dataset is available through the UK Data Service (https://ukdataservice.ac.uk) with access codes SN8346 and 5050.

## Declaration of AI use

The authors report generative AI was not used in their research or preparation of this manuscript.

