## Supplementary Materials for "Potentially modifiable mediators of the association between child abuse and dementia"

| **Variable** | **Excluded**  **N (%) / Mean (SD)** | **Included**  **N (%) / Mean (SD)** |
| --- | --- | --- |
| N | 3,362 | 5,448 |
| Dementia |  |  |
| No | 3059 (91.0) | 4935 (90.6) |
| Yes | 303 (9.0) | 513 (9.4) |
| Follow-up time | 68.6 (76.9) | 121.3 (57.4) |
| Age | 66.8 (12.4) | 65.4 (9.8) |
| Sex |  |  |
| Male | 1546 (46.0) | 2395 (44.0) |
| Female | 1816 (54.0) | 3053 (56.0) |
| Ethnicity |  |  |
| White | 3209 (95.5) | 5360 (98.4) |
| Non-white | 152 (4.5) | 88 (1.6) |
| Income quintile |  |  |
| 1 | 781 (24.7) | 800 (15.1) |
| 2 | 716 (22.6) | 941 (17.7) |
| 3 | 596 (18.8) | 1118 (21.1) |
| 4 | 561 (17.7) | 1163 (21.9) |
| 5 | 514 (16.2) | 1281 (24.2) |
| Occupation |  |  |
| Managerial and professional occupations | 957 (28.6) | 1845 (33.9) |
| Intermediate occupations | 766 (22.9) | 1420 (26.1) |
| Routine and manual occupations | 1559 (46.5) | 2123 (39.0) |
| Other | 70 (2.1) | 59 (1.1) |
| Education |  |  |
| No qualification | 1493 (44.4) | 1948 (35.8) |
| High school level | 956 (28.5) | 1681 (30.9) |
| University level | 911 (27.1) | 1818 (33.4) |
| Age finished education  No education  14 or under  15  16  17  18  19 or over  Missing | 21 (0.6)  721 (21.4)  1037 (30.8)  654 (19.5)  237 (7.0)  181 (5.4)  425 (12.6)  86 (2.6) | 14 (0.3)  826 (15.2)  1741 (32.0)  1110 (20.4)  425 (7.8)  323 (5.9)  894 (16.4)  115 (2.1) |
| Parent occupation |  |  |
| Highly skilled | 796 (23.7) | 1291 (25.5) |
| Lower skilled | 2446 (72.8) | 3890 (71.4) |
| Missing | 120 (3.6) | 167 (3.1) |
| Hypertension |  |  |
| No | 1396 (41.5) | 2096 (38.5) |
| Yes | 1966 (58.5) | 3352 (61.5) |
| Obesity |  |  |
| No | 1408 (70.7) | 3011 (71.4) |
| Yes | 583 (29.3) | 1206 (28.6) |
| Diabetes |  |  |
| No | 2985 (88.8) | 4919 (90.3) |
| Yes | 377 (11.2) | 529 (9.7) |
| Cholesterol |  |  |
| No | 1826 (54.3) | 2345 (43.0) |
| Yes | 1536 (45.7) | 3103 (57.0) |
| Nurse measured LDL |  |  |
| No | 678 (44.2) | 1356 (39.3) |
| Yes | 855 (55.8) | 2090 (60.7) |
| Anxiety |  |  |
| No | 3123 (92.9) | 5093 (93.5) |
| Yes | 239 (7.1) | 355 (6.5) |
| Depression |  |  |
| No | 2197 (65.3) | 3974 (72.9) |
| Yes | 1165 (34.7) | 1474 (27.1) |
| Smoking |  |  |
| No | 2777 (82.7) | 4726 (86.7) |
| Yes | 580 (17.3) | 722 (13.3) |
| Physical inactivity |  |  |
| No | 1026 (30.5) | 2154 (39.5) |
| Yes | 2336 (69.5) | 3294 (60.5) |
| Alcohol |  |  |
| No | 2683 (80.2) | 4208 (77.3) |
| Yes | 663 (19.8) | 1238 (22.7) |

*Supplementary Table 1. Comparisons of ELSA wave 3 core participants included in and excluded from the study.*

*.*

|  | **Sample characteristics N=5,448** | | **Complete case data N=2,874** | **Imputed data** |
| --- | --- | --- | --- | --- |
| **Variable** | **N (%) /**  **Mean (SD)** | **Missing data N (%)** | **N (%) /**  **Mean (SD)** | **% /**  **Mean (SD)** |
| **Exposure** | | | | |
| Child abuse  No  Yes | 4958 (92.7%) 88 (7.3%) | 102 (1.9%) | 2716 (94.5%) 158 (5.5%) | 92.6%  7.4% |
| **Outcome** | | | | |
| Dementia during follow-up  No  Yes | 4891 (89.8%) 557 (10.2%) | 0 (0.0%) | 2576 (89.6%) 298 (10.4%) | 89.8% 10.2% |
| Follow-up time (months) | 130.2 (65.9) | 0 (0.0%) | 132.2 (64.5) | 130.2 (65.9) |
| **Mediators** | | | | |
| **Cardiovascular health score** | 1.7 (0.9) | 1231 (22.6%) | 1.8 (0.9) | 1.6 (1.0) |
| Hypertension  No  Yes | 2096 (38.5%) 3352 (61.5%) | 0 (0.0%) | 975 (33.9%) 1899 (66.1%) | 38.5% 61.5% |
| Obesity  No  Yes | 3011 (71.4%) 1206 (28.6%) | 1231 (22.6%) | 2119 (73.6%) 755 (26.3%) | 70.6% 29.4% |
| Diabetes  No  Yes | 4919 (90.3%) 529 (9.7%) | 0 (0.0%) | 2620 (91.2%) 254 (8.8%) | 90.3% 9.7% |
| Cholesterol  No  Yes | 2345 (43.0%) 3103 (57.0%) | 0 (0.0%) | 736 (25.6%) 2138 (74.4%) | 43.0% 57.0% |
| Nurse measured LDL  No  Yes | 1356 (39.3%) 2090 (60.7%) | 2002 (36.7%) | 1109 (38.6%) 1765 (61.4%) | 53.9% 46.1% |
| **Any anxiety or depression**  **No**  **Yes** | 3927 (72.1%) 1521 (27.9%) | 0 (0.0%) | 2115 (73.6%) 759 (26.4%) | 72.1% 27.9% |
| Depression  No  Yes | 5093 (93.5%) 355 (6.5%) | 0 (0.0%) | 2142 (74.5%) 732 (25.5%) | 72.9% 27.1% |
| Anxiety  No  Yes | 3974 (72.9%) 1474 (27.1%) | 0 (0.0%) | 2683 (93.4%) 191 (6.6%) | 93.5% 6.5% |
| **Health behaviour score** | 1 (0.7) | 2 (0.0%) | 0.9 (0.7) | 1.0 (0.7) |
| Smoking  No  Yes | 4726 (86.7%) 722 (13.3%) | 0 (0.0%) | 2566 (89.3%) 308 (10.7%) | 86.7% 13.3% |
| Physical inactivity  No  Yes | 2154 (39.5%) 3294 (60.5%) | 0 (0.0%) | 1186 (41.3%) 1688 (58.7%) | 39.5% 60.5% |
| Alcohol  No  Yes | 4208 (77.3%) 1238 (22.7%) | 2 (0.0%) | 2177 (75.7%) 697 (24.3%) | 77.3% 22.7% |
| **Age finished education**  No education 14 or under 15 16 17 18 19 or over | 14 (0.3%) 826 (15.5%) 1741 (32.6%) 1110 (20.8%) 425 (8.0%) 323 (6.1%) 894 (16.8%) | 115 (2.1%) | 8 (0.3%) 430 (15.0%) 982 (34.2%) 589 (20.5%) 231 (8.0%) 158 (5.5%) 476 (16.6%) | 0.3% 15.5% 32.7% 20.8% 8.0% 6.1% 16.7% |
| **Confounders and effect modifiers** | | | | |
| Age | 66.1 (9.8) | 0 (0.0%) | 67.5 (8.7) | 66.1 (9.8) |
| Sex  Male Female | 2395 (44.0%) 3053 (56.0%) | 0 (0.0%) | 1286 (44.7%) 1588 (55.3%) | 44.0% 56.0% |
| Ethnicity  White Not White | 5360 (98.4%) 88 (1.6%) | 0 (0.0%) | 2839 (98.8%) 35 (1.2%) | 98.4% 1.6% |
| Number of books in the house at age 10  More than 10 books 10 books or fewer | 3848 (73.9%) 1357 (26.1%) | 243 (4.5%) | 2121 (73.8%) 753 (26.2%) | 73.7% 26.3% |
| Bedrooms per capita at age 10 | 0.6 (0.2) | 206 (3.8%) | 0.6 (0.2) | 0.6 (0.2) |
| Number of facilities at age 10 | 3.1 (1.4) | 195 (3.6%) | 3 (1.4) | 3.1 (1.4) |
| Parent occupation  Highly skilled  Lower skilled | 1391 (26.3%)  3890 (73.7%) | 167 (3.1%) | 763 (26.5%)  2111 (73.5%) | 26.3%  73.7% |
| **Auxiliary variables** | | | | |
| Income quintile  1 2 3 4 5 | 800 (15.1%) 941 (17.7%) 1118 (21.1%) 1163 (21.9%) 1281 (24.2%) | 145 (2.7%) | 349 (12.1%) 460 (16.0%) 612 (21.3%) 684 (23.8%) 769 (26.8%) | 15.0% 17.7% 21.1% 22.0% 24.3% |
| Occupation  Managerial and profession Intermediate occupations Routine and manual occupations Other | 1845 (33.9%) 1420 (26.1%) 2123 (39.0%)  59 (1.1%) | 1 (0.0%) | 1014 (35.3%) 775 (27.0%) 1055 (36.7%) 30 (1.0%) | 33.9% 26.1% 39.0%  1.1% |
| Education  No qualification High school level University level | 1948 (35.8%) 1681 (30.9%) 1818 (33.4%) | 1 (0.0%) | 1005 (35.0%) 888 (30.9%) 981 (34.1%) | 35.8% 30.9% 33.4% |

*Supplementary Table 2. Sample characteristics.*

|  | **Child abuse** | | **Dementia** | |
| --- | --- | --- | --- | --- |
| **Variable** | **No (92.6%)**  **% / Mean (SD)** | **Yes (7.4%)**  **% / Mean (SD)** | **No (89.8%)**  **% / Mean (SD)** | **Yes (10.2%)**  **% / Mean (SD)** |
| **Exposure** | | | | |
| Child abuse  No  Yes |  |  | 92.6% 7.4% | 92.0% 8.0% |
| **Outcome** | | | | |
| Dementia during follow-up  No  Yes | 89.8% 10.2% | 89.1% 10.9% |  |  |
| Follow-up time (months) | 129.4 (65.9) | 139.7 (64.7) | 135.2 (65.2) | 86.4 (54.8) |
| **Mediators** | | | | |
| **Cardiovascular health score** | 1.56 (1.0) | 1.5 (1.1) | 1.6 (1.0) | 1.8 (0.9) |
| Hypertension  No  Yes | 37.9% 62.1% | 45.9% 54.1% | 39.8% 60.2% | 26.4% 73.6% |
| Obesity  No  Yes | 71.0% 29.0% | 65.2% 34.8% | 70.8% 29.2% | 68.7% 31.3% |
| Diabetes  No  Yes | 90.3% 9.7% | 90.7% 9.3% | 90.7% 9.3% | 86.5% 13.5% |
| Cholesterol  No  Yes | 42.7% 57.3% | 47.5% 52.5% | 43.4% 56.6% | 39.9% 60.1% |
| Nurse measured LDL  No  Yes | 53.4% 46.6% | 57.5% 42.5% | 53.8% 46.2% | 52.7% 47.3% |
| **Any anxiety or depression**  **No**  **Yes** | 73.4% 26.6% | 55.2% 44.8% | 72.8% 27.2% | 65.5% 34.5% |
| Depression  No  Yes | 74.3% 25.7% | 56.0% 44.0% | 73.7% 26.3% | 66.2% 33.8% |
| Anxiety  No  Yes | 94.3% 5.7% | 83.2% 16.8% | 93.6% 6.4% | 92.6% 7.4% |
| **Health behaviour score** | 1.0 (0.7) | 1.0 (0.7) | 1.0 (0.7) | 1.0 (0.7) |
| Smoking  No  Yes | 87.2% 12.8% | 80.9% 19.1% | 86.4% 13.6% | 90.1% 9.9% |
| Physical inactivity  No  Yes | 39.6% 60.4% | 38.7% 61.3% | 40.8% 59.2% | 28.5% 71.5% |
| Alcohol  No  Yes | 77.0% 23.0% | 80.9% 19.1% | 77.2% 22.8% | 78.1% 21.9% |
| **Age finished education**  No education 14 or under 15 16 17 18 19 or over | 0.3% 16.1% 32.5% 20.6% 7.9% 6.0% 16.6% | 0.0% 8.1% 34.4% 23.4% 8.6% 7.0% 18.5% | 0.2% 14.2% 33.1% 21.2% 7.8% 6.3% 17.2% | 0.6% 27.1% 29.1% 17.1% 9.6% 4.3% 12.3% |
| **Confounders and effect modifiers** | | | | |
| Age | 66.5 (9.8) | 61.8 (8.4) | 65.3 (9.5) | 73.3 (8.9) |
| Sex  Male Female | 44.6% 55.4% | 36.3% 63.7% | 43.7% 56.3% | 46.5% 53.5% |
| Ethnicity  White Not White | 98.4% 1.6% | 98.5% 1.5% | 98.4% 1.6% | 98.6% 1.4% |
| Number of books in the house age 10  More than 10 books None or very few (0-10 books) | 73.8%  26.2% | 73.2%  26.8% | 74.7%  25.3% | 65.1%  34.9% |
| Bedrooms per capita age 10 | 0.6 (0.2) | 0.6 (0.2) | 0.6 (0.2) | 0.6 (0.2) |
| Number of facilities age 10 | 3.1 (1.4) | 3.4 (1.3) | 3.1 (1.4) | 2.8 (1.5) |
| Parent occupation  Highly skilled  Lower skilled | 26.0%  74.0% | 29.5%  70.% | 26.4%  73.6% | 25.8%  74.2% |
| **Auxiliary variables** | | | | |
| Income quintile  1 2 3 4 5 | 14.7% 17.5% 21.1% 22.3% 24.5% | 18.2% 20.4% 20.7% 18.6% 22.1% | 14.5% 17.6% 21.0% 22.1% 24.7% | 18.9% 18.7% 21.6% 20.5% 20.2% |
| Occupation  Managerial and profession  Intermediate occupations  Routine and manual occupations  Other | 33.8%  26.2%  38.9%  1.1% | 34.4%  24.7% 39.6%  1.3% | 34.3%  26.0% 38.6%  1.0% | 30.2%  26.4% 41.8%  1.6% |
| Education  No qualification High school level University level | 36.2% 30.9% 32.9% | 30.0% 30.5% 39.6% | 35.2% 30.8% 34.0% | 40.9% 31.4% 27.6% |

*Supplementary Table 3. Sample characteristics of imputed data cross-tabulated by child abuse and dementia.*

|  |  |  | **Associations between child abuse and mediators** | | | **Associations between mediators and dementia** | | |
| --- | --- | --- | --- | --- | --- | --- | --- | --- |
| **Analysis** | **Category** | **Mediator** | **OR/β** | **95% CI** | | **HR** | **95% CI** | |
| Primary analysis | Cardiovascular Health | Hypertension (OR) | 1.00 | 0.81 | 1.25 | 1.15 | 0.95 | 1.40 |
|  |  | Obesity (OR) | 1.20 | 0.91 | 1.59 | 1.31 | 1.07 | 1.60 |
|  |  | Cholesterol (OR) | 0.99 | 0.80 | 1.22 | 0.88 | 0.74 | 1.05 |
|  |  | Diabetes (OR) | 1.17 | 0.82 | 1.67 | 1.45 | 1.13 | 1.85 |
|  |  | Cardiovascular score (β) | 0.05 | -0.06 | 0.16 | 1.11 | 1.01 | 1.22 |
|  | Mental Health | Depression (OR) | 2.24 | 1.81 | 2.77 | 1.58 | 1.32 | 1.90 |
|  |  | Anxiety (OR) | 2.90 | 2.16 | 3.89 | 1.58 | 1.15 | 2.19 |
|  |  | Depression or anxiety (OR) | 2.20 | 1.78 | 2.72 | 1.56 | 1.30 | 1.87 |
|  | Health Behaviour | Smoking (OR) | 1.31 | 1.00 | 1.72 | 1.35 | 1.02 | 1.80 |
|  |  | Physical inactivity (OR) | 1.20 | 0.97 | 1.49 | 1.43 | 1.18 | 1.74 |
|  |  | Alcohol (OR) | 0.83 | 0.64 | 1.08 | 0.88 | 0.71 | 1.08 |
|  |  | Health behaviour score (β) | 0.05 | -0.02 | 0.12 | 1.18 | 1.04 | 1.34 |
|  | Education | Education (β) | -0.01 | -0.16 | 0.14 | 1.01 | 0.95 | 1.07 |
| Not adjusting for childhood SES | Cardiovascular Health | Hypertension (OR) | 1.00 | 0.81 | 1.25 | 1.16 | 0.96 | 1.41 |
|  |  | Obesity (OR) | 1.20 | 0.91 | 1.58 | 1.33 | 1.09 | 1.63 |
|  |  | Cholesterol (OR) | 0.98 | 0.79 | 1.22 | 0.86 | 0.73 | 1.03 |
|  |  | Diabetes (OR) | 1.16 | 0.81 | 1.66 | 1.48 | 1.16 | 1.89 |
|  |  | Cardiovascular score (β) | 0.05 | -0.06 | 0.15 | 1.12 | 1.02 | 1.23 |
|  | Mental Health | Depression (OR) | 2.25 | 1.82 | 2.79 | 1.64 | 1.37 | 1.96 |
|  |  | Anxiety (OR) | 2.94 | 2.20 | 3.94 | 1.64 | 1.19 | 2.26 |
|  |  | Depression or anxiety (OR) | 2.22 | 1.79 | 2.74 | 1.61 | 1.35 | 1.93 |
|  | Health Behaviour | Smoking (OR) | 1.32 | 1.01 | 1.73 | 1.45 | 1.09 | 1.92 |
|  |  | Physical inactivity (OR) | 1.21 | 0.97 | 1.49 | 1.47 | 1.22 | 1.78 |
|  |  | Alcohol (OR) | 0.84 | 0.65 | 1.09 | 0.84 | 0.69 | 1.03 |
|  |  | Health behaviour score (β) | 0.05 | -0.02 | 0.12 | 1.19 | 1.05 | 1.34 |
|  | Education | Education (β) | -0.03 | -0.19 | 0.12 | 1.05 | 0.99 | 1.10 |
| Complete case | Cardiovascular Health | Hypertension (OR) | 0.89 | 0.63 | 1.24 | 1.35 | 1.02 | 1.78 |
|  |  | Obesity (OR) | 1.31 | 0.92 | 1.85 | 1.51 | 1.18 | 1.94 |
|  |  | Cholesterol (OR) | 1.08 | 0.75 | 1.61 | 1.00 | 0.76 | 1.30 |
|  |  | Diabetes (OR) | 1.57 | 0.91 | 2.58 | 1.22 | 0.84 | 1.77 |
|  |  | Cardiovascular score (β) | 0.08 | -0.07 | 0.23 | 1.24 | 1.08 | 1.42 |
|  | Mental Health | Depression (OR) | 2.13 | 1.52 | 2.99 | 1.52 | 1.17 | 1.96 |
|  |  | Anxiety (OR) | 3.13 | 1.99 | 4.81 | 1.21 | 0.73 | 2.03 |
|  |  | Depression or anxiety (OR) | 2.06 | 1.47 | 2.89 | 1.50 | 1.16 | 1.93 |
|  | Health Behaviour | Smoking (OR) | 1.61 | 1.02 | 2.47 | 1.60 | 1.07 | 2.39 |
|  |  | Physical inactivity (OR) | 1.46 | 1.04 | 2.07 | 1.34 | 1.04 | 1.72 |
|  |  | Alcohol (OR) | 0.88 | 0.58 | 1.30 | 0.78 | 0.59 | 1.05 |
|  |  | Health behaviour score (β) | 0.12 | 0.01 | 0.23 | 1.13 | 0.95 | 1.34 |
|  | Education | Education (β) | -0.04 | -0.27 | 0.19 | 1.00 | 0.93 | 1.09 |

*Supplementary Table 4. Regression modelling of the association between child abuse, mediators and dementia.*

*Note: Logistic regression of mediator in association with child abuse. OR presented for binary mediators and β coefficient presented for continuous mediators. Cox-proportional hazards model of dementia in association with mediator. HR presented for all mediators.*

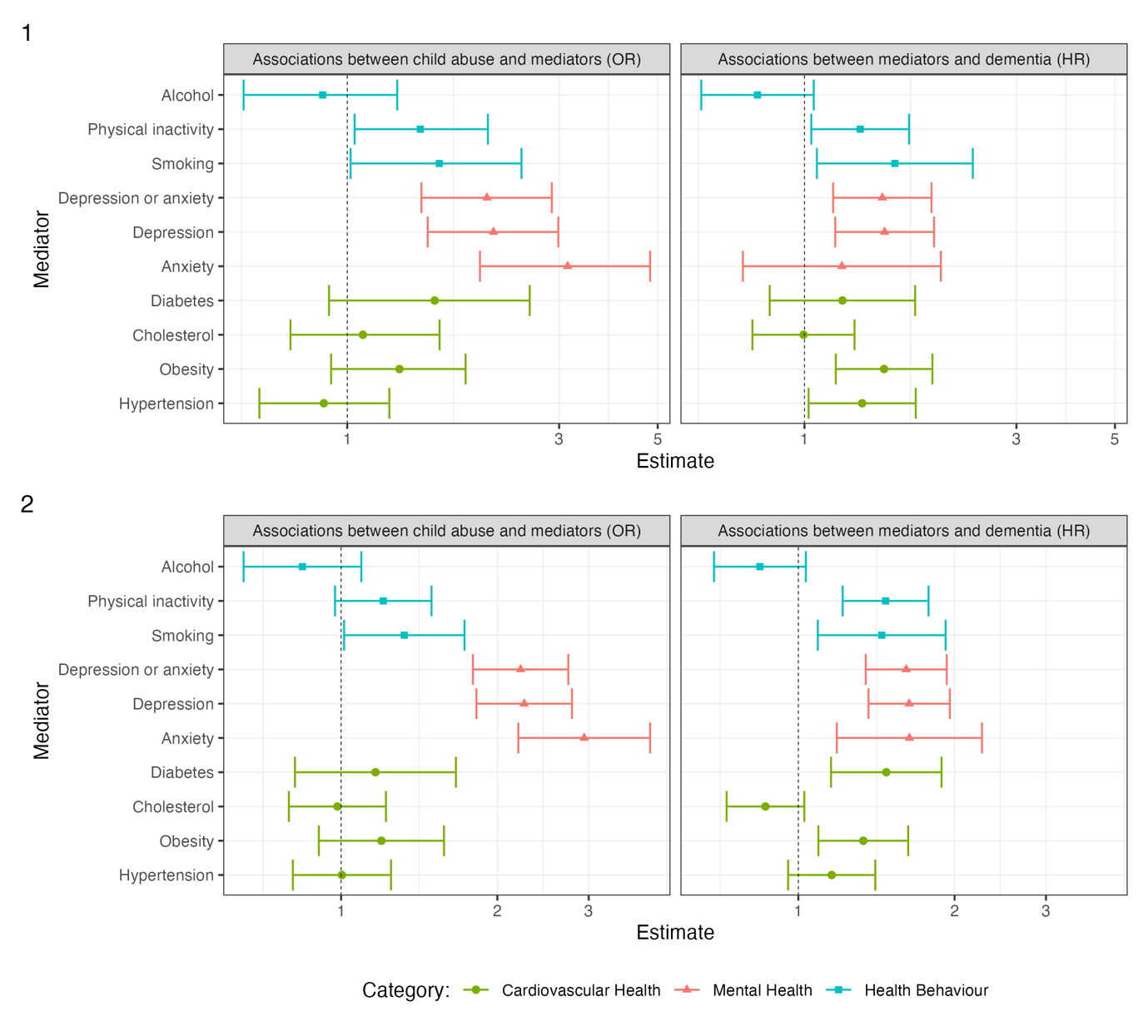

*Supplementary Figure 1. Sensitivity analysis of regression models of the associations between child abuse, binary meditators and dementia incidence. 1) Complete case analysis. 2) Analysis not adjusting for childhood SES.*

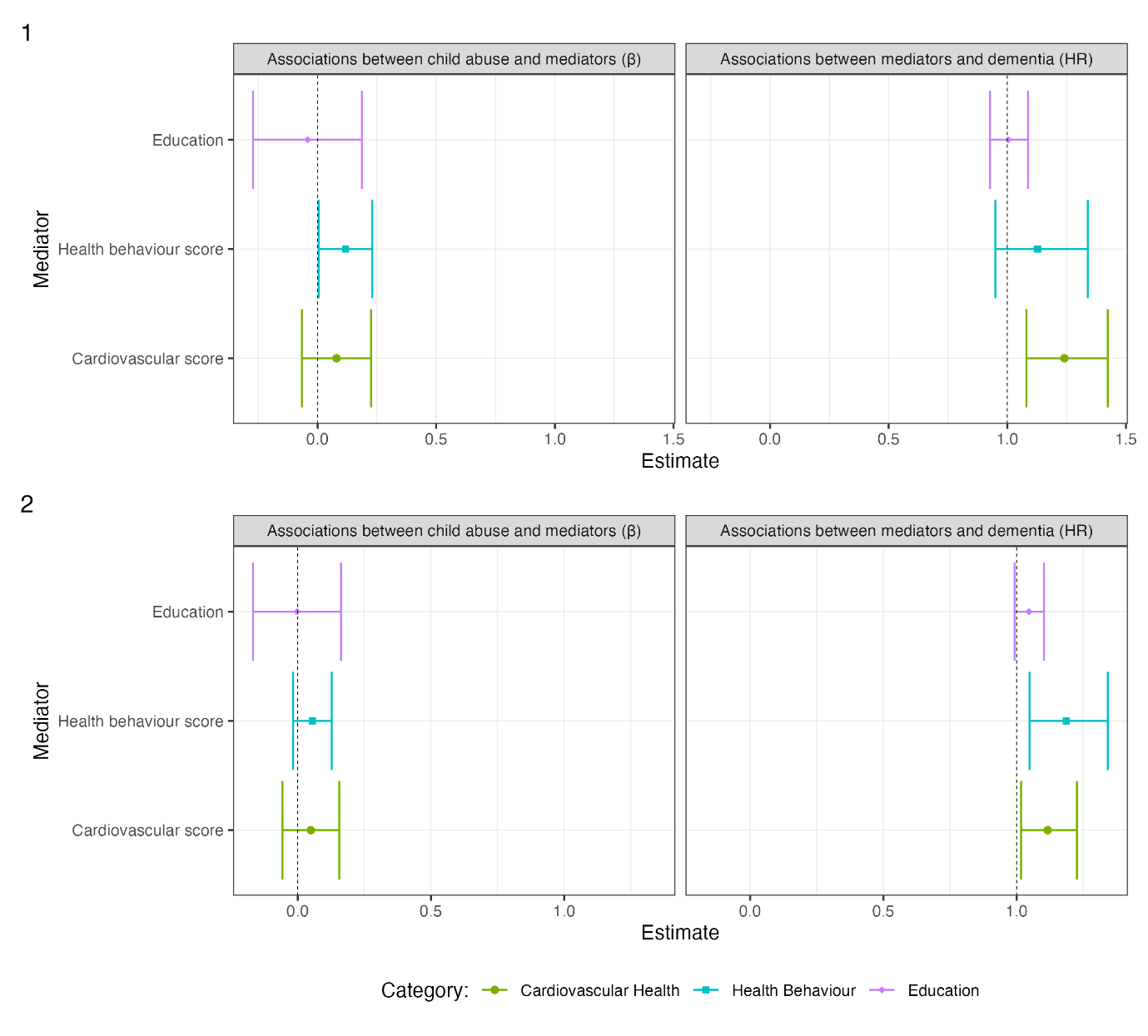

*Supplementary Figure 2. Sensitivity analysis of regression models of the associations between child abuse, continuous meditators and dementia incidence. 1) Complete case analysis. 2) Analysis not adjusting for childhood SES.*

|  | **Associations between child abuse and mediators** | | | **Associations between mediators and dementia** | | |
| --- | --- | --- | --- | --- | --- | --- |
| **Analysis** | **OR** | **95% CI** | | **HR** | **95% CI** | |
| Primary analysis | 0.98 | 0.93 | 1.04 | 0.91 | 0.76 | 1.09 |
| Not adjusting for childhood SES | 0.98 | 0.93 | 1.03 | 0.89 | 0.74 | 1.06 |
| Complete case | 0.90 | 0.65 | 1.27 | 1.04 | 0.82 | 1.32 |

*Supplementary Table 5. Sensitivity analysis considering nurse measured LDL only as a measure of high cholesterol.*

| **Analysis** | **Education definition** | **IPW** | **OR/β** | **95% CI** | |
| --- | --- | --- | --- | --- | --- |
| No exclusions | Continuous (β) | No | -0.16 | 0.14 | -0.01 |
|  |  | Yes | -0.08 | -0.25 | 0.09 |
|  | Binary - 15 under (OR) | No | 0.96 | 0.76 | 1.21 |
|  |  | Yes | 0.93 | 0.73 | 1.18 |
|  | Binary - 14 under (OR) | No | 0.95 | 0.60 | 1.50 |
|  |  | Yes | 0.88 | 0.57 | 1.36 |
| No SES adjustment | Continuous (β) | No | 0.00 | -0.17 | 0.16 |
|  |  | Yes | -0.05 | -0.22 | 0.12 |
|  | Binary - 15 under (OR) | No | 0.98 | 0.79 | 1.21 |
|  |  | Yes | 0.96 | 0.76 | 1.22 |
|  | Binary - 14 under (OR) | No | 0.93 | 0.60 | 1.44 |
|  |  | Yes | 0.92 | 0.60 | 1.42 |
| Exclude those with no education | Continuous (β) | No | 0.00 | -0.14 | 0.15 |
|  |  | Yes | -0.07 | -0.24 | 0.10 |
|  | Binary - 15 under (OR) | No | 0.97 | 0.76 | 1.23 |
|  |  | Yes | 0.93 | 0.73 | 1.19 |
|  | Binary - 14 under (OR) | No | 0.97 | 0.61 | 1.54 |
|  |  | Yes | 0.90 | 0.58 | 1.39 |
| Exclude abuse after education end | Continuous (β) | No | -0.06 | -0.25 | 0.13 |
|  |  | Yes | -0.04 | -0.27 | 0.18 |
|  | Binary - 15 under (OR) | No | 0.89 | 0.66 | 1.21 |
|  |  | Yes | 0.92 | 0.59 | 1.44 |
|  | Binary - 14 under (OR) | No | 0.97 | 0.51 | 1.82 |
|  |  | Yes | 1.23 | 0.49 | 3.07 |

*Supplementary Table 6. Sensitivity analysis of the association between child abuse and age leaving education.*

|  | **Primary analysis** | | | **Mental health only** | | | **Mental health only, adjusting for other mediators** | | | **Not adjusting for childhood SES** | | | **Complete case** | | |
| --- | --- | --- | --- | --- | --- | --- | --- | --- | --- | --- | --- | --- | --- | --- | --- |
| **Effect** | **HR** | **95% CI** | | **HR** | **95% CI** | | **HR** | **95% CI** | | **HR** | **95% CI** | | **HR** | **95% CI** | |
| Rcde | 1.66 | 1.12 | 1.65 | 1.65 | 1.11 | 2.19 | 1.66 | 1.12 | 2.20 | 1.71 | 1.16 | 2.26 | 1.35 | 0.74 | 2.09 |
| Rpnde | 1.66 | 1.12 | 1.65 | 1.65 | 1.11 | 2.19 | 1.66 | 1.12 | 2.20 | 1.71 | 1.16 | 2.26 | 1.35 | 0.74 | 2.09 |
| Rtnde | 1.66 | 1.12 | 1.65 | 1.65 | 1.11 | 2.19 | 1.66 | 1.12 | 2.20 | 1.71 | 1.16 | 2.26 | 1.35 | 0.74 | 2.09 |
| Rpnie | 1.08 | 1.03 | 1.08 | 1.08 | 1.03 | 1.13 | 1.07 | 1.03 | 1.12 | 1.09 | 1.04 | 1.15 | 1.09 | 1.02 | 1.18 |
| Rtnie | 1.08 | 1.03 | 1.08 | 1.08 | 1.03 | 1.13 | 1.07 | 1.03 | 1.12 | 1.09 | 1.04 | 1.15 | 1.09 | 1.02 | 1.18 |
| Rte | 1.80 | 1.21 | 1.78 | 1.78 | 1.20 | 2.36 | 1.78 | 1.20 | 2.35 | 1.87 | 1.27 | 2.47 | 1.46 | 0.78 | 2.29 |
| Proportion Mediated | 0.18 |  | 0.17 | 0.17 | 0.17 |  | 0.15 |  |  | 0.18 |  |  | 0.25 |  |  |

*Supplementary Table 7. Interventional effects for mediation analysis.*

*Note: Rcde: Randomised controlled direct effect. Rpnde: Randomised pure natural direct effect. Rtnde: Randomised true natural direct effect. Rpnie: Randomised pure natural indirect effect. Rtnie: Randomised true natural indirect effect. Rte: Randomised total effect*

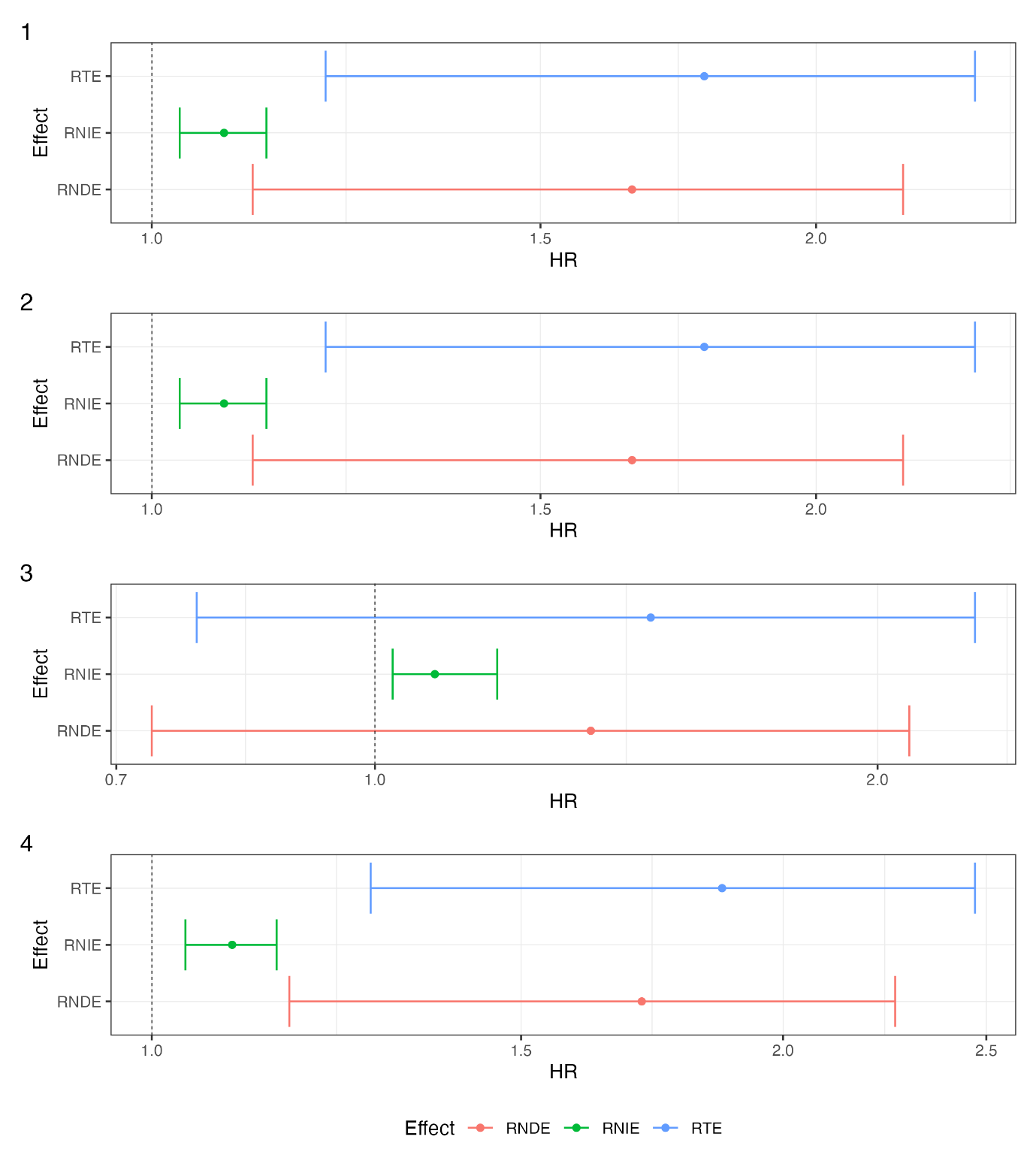
*Supplementary Figure 3. Interventional effects for mediation analysis. 1) Considering only mental health as a mediator. 2) Considering only mental health as a mediator, adjusting for other mediators as confounders. 3) Complete case analysis. 4) Not adjusting for childhood SES.*

*Note: Joint mediation effects of education, health behaviours, cardiovascular health and mental health on the association between child abuse and dementia incidence. RNDE: Randomised natural direct effect. RNIE: Randomised natural indirect effect. RTE: Randomised total effect.*
